# Evaluating the use of a novel low-cost measurement insole to characterise plantar foot strain during gait loading regimes

**DOI:** 10.1101/2023.04.04.23288138

**Authors:** Sarah R Crossland, Heidi J Siddle, Claire L Brockett, Peter Culmer

## Abstract

Under plantar loading regimes, it is accepted that both pressure and shear strain biomechanically contribute to formation and deterioration of diabetic foot ulceration (DFU). Plantar foot strain characteristics in the at-risk diabetic foot are little researched due to lack of measurement devices. Plantar pressure comparatively, is widely quantified and used in the characterisation of diabetic foot ulceration risk, with a range of clinically implemented pressure measurement devices on the market. With the development of novel strain quantification methods in its infancy, feasibility testing and validation of these measurement devices for use is required. Initial studies centre on normal walking speed, reflecting common activities of daily living, but evaluating response to differing gait loading regimes is needed to support the use of such technologies for potential clinical translation. This study evaluates the effects of speed and inclination on stance time, strain location and strain response using a low-cost novel strain measurement insole. The STrain Analysis and Mapping of the Plantar Aspect (STAMPS) insole has been developed, and feasibility tested under self-selected normal walking speeds to characterise plantar foot strain, with testing beyond this limited regime required. A treadmill was implemented to standardise speed and inclination for a range of daily plantar loading conditions. A small cohort, comprising of five non-diabetic participants, were examined at slow (0.75 m/s), normal (1.25 m/s) and brisk (2 m/s) walking speeds and normal speed at inclination (10% gradient). Plantar strain active regions were seen to increase with increasing speed across all participants. With inclination, it was seen that strain active regions reduce in the hindfoot and show a tendency to forefoot with discretionary changes to strain seen. Stance time decreases with increasing speed, as expected, with reduced stance time with inclination. Comparison of the strain response and stance time should be considered when evaluating foot biomechanics in diabetic populations to assess strain time interval effects. This study supports the evaluation of the STAMPS insole to successfully track strain changes under differing plantar loading conditions and warrants further investigation of healthy and diabetic cohorts to assess the implications for use as a risk assessment tool for DFU.

## 1 INTRODUCTION

The global diabetic population has increased significantly in recent decades with growth predicted to continue (International Diabetes Federation, 2019). With this comes a rise in the associated development of diabetic foot disease. From this population it is expected up to 25% will develop diabetic foot ulceration (DFU) within their lifetime (Armstrong et al., 2017). The associated healing times and treatment pathway requirements for DFU lead to a labour and cost intensive process with over £900 million spent annually in the UK market alone (Kerr et al., 2019), which is neither beneficial to the patient or healthcare provider. Prophylactic intervention is fundamental to reducing DFU rates, but is often unsupported in clinical practice due in part to poor evidence base and cost to implement across the at-risk diabetic population (Heuch and Streak Gomersall, 2016; Kerr et al., 2019; Bus et al., 2020). The current evidence base for orthotic intervention is focused on pressure as a predictor of ulceration risk to inform offloading (Bus et al., 2020). This has centered the development of diabetic foot risk assessment tools to solely focus on pressure. While elevated and sustained plantar pressures in DFU are well researched, there is often discrepancy between ulcer location and the peak plantar pressure site (Lavery et al., 2003). Shear stress on the foot is thought in part to contribute to this deviation in expected location (Jones et al., 2022), but remains little understood and is not measured in risk assessment of the diabetic foot due to the poor availability of measurement tools.

The complexities seen in the feet of people with diabetes leads to a requirement of bespoke treatment approaches. This in turn drives the development of objective risk assessment tools that allow quantifiable metrics of the at-risk diabetic foot and allow for earlier prophylactic interventions to reduce DFU formation risk and work towards preventing long term escalation of treatment costs (Bus et al., 2020). Current approaches to quantify shear at the plantar surface utilise a wide range of technologies including capacitive sensors and strain gauges (Rajala and Lekkala, 2014), but have not established a clinically viable tool (Yavuz et al., 2007; Jones et al., 2022). With a gulf in technology addressing both the pressure and shear components of plantar load.

Whilst pressure time integral is considered alongside peak plantar pressure and average pressure in assessing DFU risk (Keijsers et al., 2010; Waaijman and Bus, 2012; Bus and Waaijman, 2013), the contribution of shear strain time integral remains unclear, due to the limited systems available to measure strain in lieu of shear forces and no current clinically utilised techniques for data collection. Yavuz et al. (2008) employed a custom built sensor platform to measure normal and tangential forces simultaneously of the unshod foot during stance phase to derive pressure and shear time integrals for a diabetic and non-diabetic cohort. This showed by an increase in both time integrals for the diabetic population and led to calls for further investigation of temporal strain responses.

The current pressure data capture techniques are divided into two distinct focuses of shod or unshod measures. Whilst unshod measures can give an understanding of intrinsic pressures due to anatomical variances and gait deviations, they do not reflect the activities of daily living where footwear is worn. However, in clinic these pressure devices, including pressure plates (Abdul Razak et al., 2012), offer a convenient method of data capture with which to inform orthoses design. Shod pressure data allows data to be collected during these activities of daily living to provide a representative understanding of the pressure events acting upon the diabetic foot. Technologies including as pedar® [Novel GmbH, Munchen Germany] pressure measurement insoles are currently used in clinical and research settings to achieve shod pressure data collection. Recent trends include the emerging market of pressure reporting insoles offering real-time feedback to inform user behaviour and minimise DFU risk(Chatwin et al., 2021). For both of these methods, the cost, initial set-up, calibration requirements and training are prohibitive factors to their implementation in a clinical environment.

The shod environment also presents influential factors which may instigate the formation of ulceration due to pressure and shear events leading to mechanical tissue stress (Lord and Hosein, 2000). The interfaces between the foot, sock and shoe must be considered in this instance, alongside the pressure changes brought about by the footwear design and the influence on tissue stress (van Netten et al., 2018). To begin to understand the effect on differing loading regimes to the plantar aspect of the foot within the shod environment, controlled speed and inclination trials have been employed (Segal et al., 2004; Kernozek et al., 1996; Warren et al., 2004; Ho et al., 2010) using the pedar® pressure measurement insole. This method allows for a benchmark to be provided, allowing reporting of patterns in pressure deviation with changing speeds that reflect activities of daily living.

Current clinical pressure measurements systems, such as pedar® [Novel GmbH, Munchen Germany], provide the functionality to monitor pressure response changes under differing loading regimes in the feet of people with diabetes. Recognition of the need to assess the plantar aspect during functional gait is seen with use of technologies such as pedar® and should form a basis for future monitoring method requirements. This clinical need for a loading regime responsive assessment method drove the methodology to analyse the STAMPS insole response (Crossland et al., 2023).

The development of the STrain Analysis and Mapping of the Plantar Surface (STAMPS) insole by Crossland et al. (2023) bridges these gaps in the literature by allowing for strain assessment as a surrogate for the components of plantar load during gait. Digital image correlation (DIC), computer vision tracking of changes to an applied stochastic speckle pattern (Michael A. et al., 2009), is used here to quantify the cumulative effects of plantar loading in the form of strain imparted on a plastically deformable insole during gait. Currently STAMPS has been optimised for functionality and feasibility tested at self-selected normal walking speeds and without a gradient. This paper uses the STAMPS insole technique as a responsive tool to evaluate changes in strain characteristics aligned to changes in walking speed and inclination, including stance time, strain location and strain response.

## 2 MATERIALS AND METHODS

### 2.1 Study Protocol

To provide a consistent achieved walking speed and inclination across all studies a Nordictrack C200 Treadmill was implemented for use. Trials were selected to be conducted at 0.75 m/s, 1.25 m/s and 2 m/s speeds to reflect a slow, lower bound normal and brisk walking pace. These values align with conducted treadmill trials to monitor pressure variance with speed during gait using pedar ® (Segal et al., 2004) and also reflect the range of speeds that might be adopted in typical activities of daily living. Inclination was set to a gradient of 10% reflecting a mid value condition selected by Ho et al. (2010). It was decided that for the purpose of this study, the inclination trial would deviate from Ho et al. (2010) and be conducted at the ‘normal’ 1.25 m/s speed, to reflect the expected general gait reported in a slower population (Segal et al., 2004), such as may be expected in the ageing diabetic population. Due to safety limitations, the treadmill belt restricts starting at the target speed and instead provides an acceleration to reach this speed. The treadmill acceleration profiles were collated using image analysis, recorded using a Nikon D5300 with AF-S Nikkor Lens (Nikon), to track belt speed changes under the three speed conditions (Padulo et al., 2014).

#### 2.1.1 Insole Manufacture

The STAMPS plastically deformable insoles were prepared following the protocol described previously (Crossland et al., 2023). A commercial clay roller (CT-500, North Star Polaris) was used to provide a targeted 5 mm thickness plasticine slab from which the insoles were cut to size requirements. Cross patterned Nylon mesh was used to reinforce the base of the insole and provide a posterior tab for ease of removal following use (Fig. 1). The optimised computer generated stochastic speckle (Correlated Solutions Speckle Generator, v1.0.5), consisting of a 0.8 mm speckle with a 65% pattern density and 75% pattern variation, was applied via a thin film, 180 *µ*m, temporary tattoo (Silhoutte, USA) for the purpose of DIC. The insoles were allowed to rest for a period of 24 hours minimum prior to use after moulding to allow for any temporal hardening effects (Chijiiwa et al., 1981). The insoles were then stored at a controlled 15 °temperature prior to use, in line with Crossland et al. (2023) findings on storage and use optimisation for ten step gait studies.

**Figure 1.**
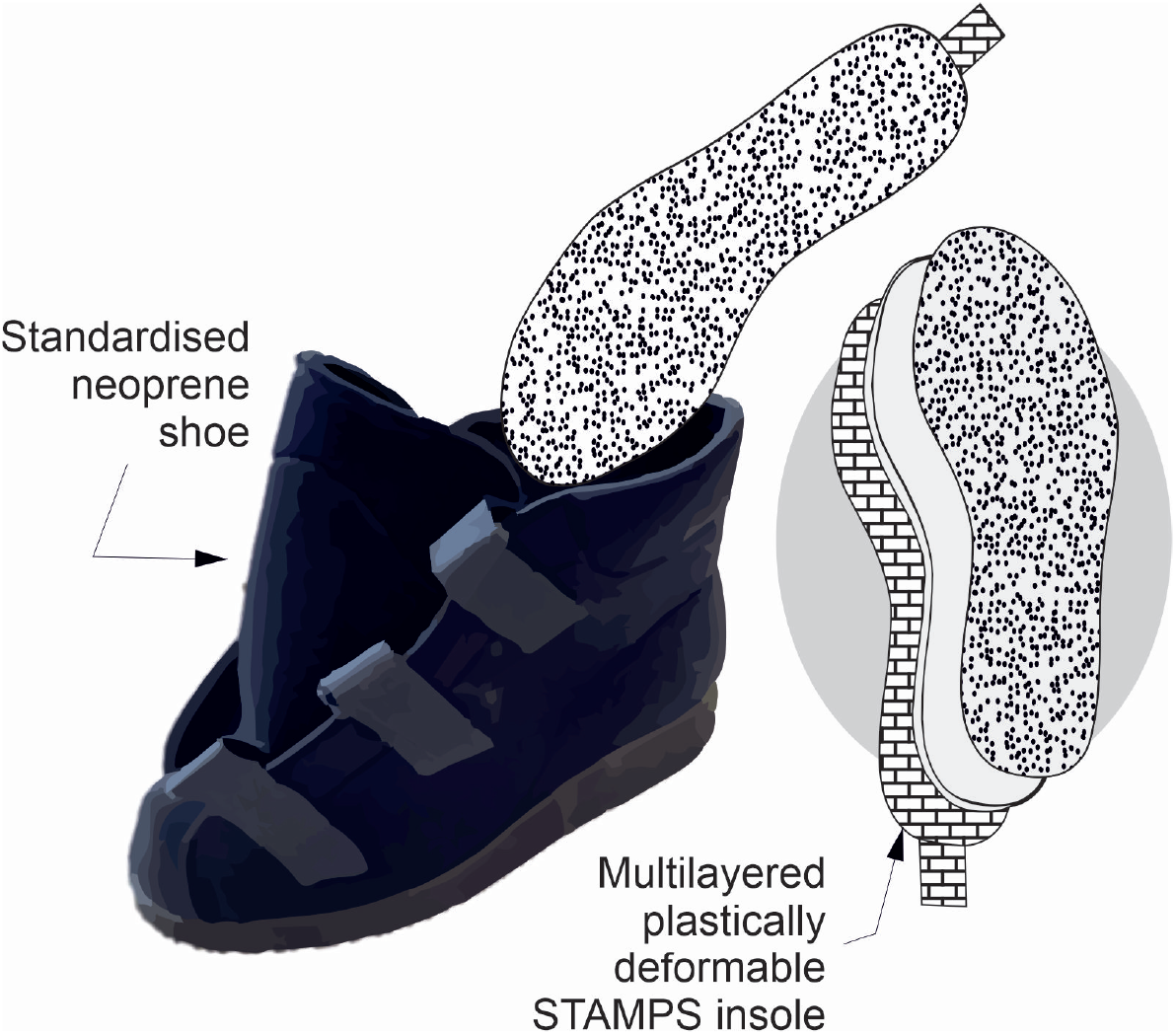
STAMPS insole layer view schematic showing standardised footwear utilised within the participant study.

#### 2.1.2 Participant Study

To verify the proposed study protocol, a participant cohort was recruited. The aim of which was to assess the ability of the STAMPS insole to effectively detect strain changes under differing loading regimes through controlled speed and inclination trials. A five participant non-diabetic cohort was recruited and provided consent, see Table 1. The University of Leeds Engineering and Physical Sciences joint Faculty Research Ethics Committee granted ethics approval (LTMECH-005) for the study design. The study assessed right foot stance phase loading solely, with each participant provided with a STAMPS insole for the right footwear with a contra-lateral sham insole in the left footwear to reduce inconsistency in leg length. Standardised neoprene footwear (Ninewells Boot, Chaneco LTD) were used for consistency across all participants. Participants were asked to walk for ten steps on the right foot during each trial, inline with insole usability limits procured from insole optimisation (Crossland et al., 2023). Three repeats were taken at each trialled speed and at inclination. Images were recorded of the STAMPS insole before and after undertaking each trial and participants were recorded using an camera recording at 50 fps (Nikon D5300, Nikon) to capture stance phase contact time.

**Table 1.** Participant characterisation data collated for speed and inclination treadmill study.

| Participant | Gender | Height (m) | Weight (kg) | Age Range (Years) | Shoe Size (UK) |
| --- | --- | --- | --- | --- | --- |
| 1 | F | 1.75 | 64.5 | 26-30 | 7 |
| 2 | M | 1.94 | 83.2 | 31-35 | 12 |
| 3 | M | 1.85 | 85.0 | 26-30 | 11 |
| 4 | M | 1.90 | 77.6 | 26-30 | 12 |
| 5 | M | 1.82 | 83.1 | 26-30 | 11 |

### 2.2 Plantar Strain Analysis

Commercially available DIC software (GOM Correlate 2019) was used for first stage post image DIC analysis to allow for the generation as insole strain maps. Strains were determined relative to the reference photo of the insole taken prior to each trial. For exportation of the data for post processing to derive positional strain values, an equidistant spread of points at 6.5 mm intervals was applied to each insole.

Post processing was conducted in MATLAB (R2021b) for implementation of custom scripts to improve visualisation and allow for anatomical regional analysis of strain data. Pedar® [Novel GmbH, Munchen Germany] used as a tool for risk assessing the diabetic foot, employs an Automask feature to divide the foot by regions of anatomically significance for segmented analysis in areas of DFU prevalence (Pit’hová et al., 2007). Replicative masking across key anatomical landmarks was applied to the post-processed strain maps and aligned anatomical by a qualified orthotist (SRC). A reductive masking approach was then used to combine localised regions which would be difficult to distinguish clearly through assessment of the insole imaging. A resulting eight region mask was applied to determine strain outputs (Fig. 2), covering: hallux, second to fifth toes, first metatarsal head, second and third metatarsal head, fourth and fifth metatarsal heads, lateral midfoot, medial midfoot and calcaneus. Average and peak strains across each segment were determined.

**Figure 2.**
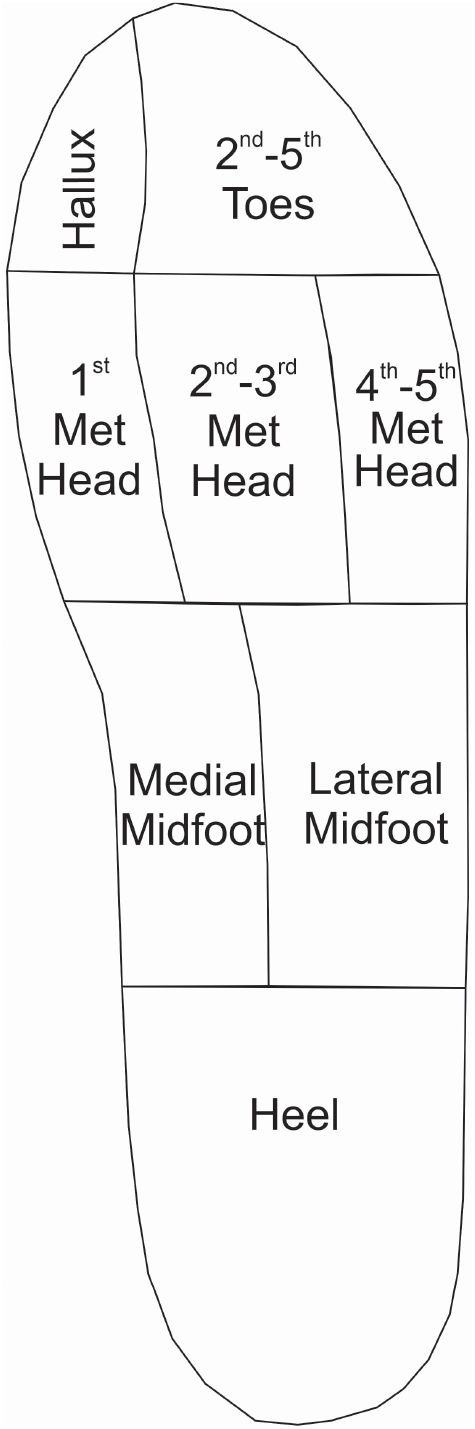
Anatomically defined regional mask which is used to catergorise strain output data for each participant trial.

## 3 RESULTS

All trials were successfully completed for ten stance phases on the right foot by each participant. Fig.3 provides representative strain visualisation outputs from a single participant, showing the three repeated trials under each loading regime. The figure shows regions identified as being strain active increase with increasing speed, strain within the active regions also increases in line with the increasing speed. Fig. 3 also highlights the variance between the two trials conducted at 1.25 m/s at 0% and 10% inclinations. Strain active regions are maintained in the forefoot with a reduction in activity seen in the hindfoot with increased inclination. These patterns are seen generally across all participants, with supplementary corresponding figures supplied for each of the remaining participants.

**Figure 3.**
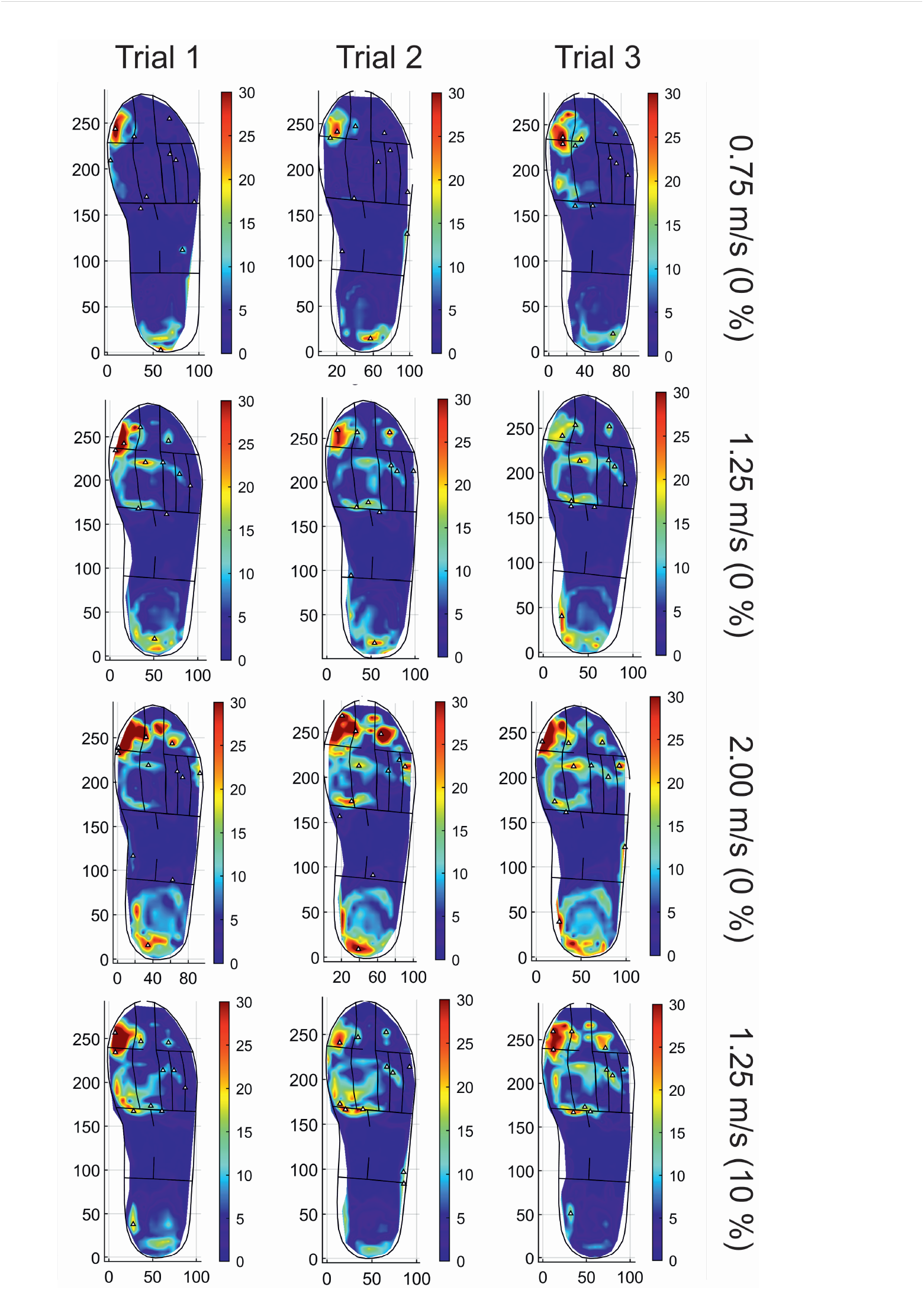
A representative example of strain profiles for repeated trialled speeds and inclinations for one participant (P03).

Tables 2 and 3 show the averaged trial strains, standard deviations and percentage strain changes seen between speed changes (Table 2) and due to inclination change (Table 3). The trend between increasing speed and increasing average and peak strain can be seen for all participants in the majority of anatomical regions. There is some variance in the reported strain changes for inclination across differing anatomical regions and participants. All participants show a reduction in strain with increasing inclination at the rearfoot, in line with the reduction in strain active regions as seen in Fig. 3.

**Table 2.**
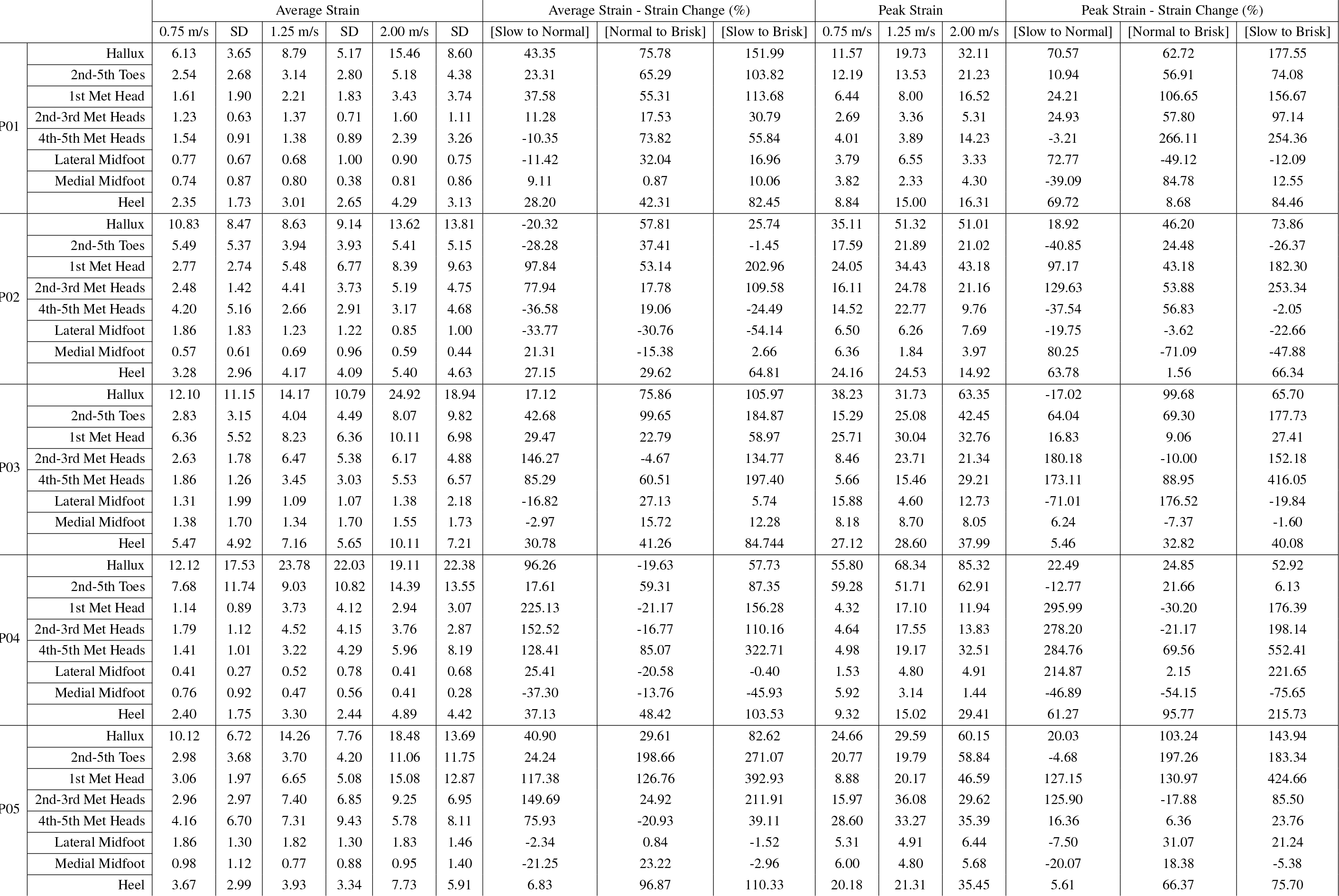
Anatomical regional average and peak strains to 2 d.p. and standard deviations (SD) averaged across all three repeat trials, with comparative percentage strain changes for all participants (P01-P05) at walking speeds of 0.75, 1.25 and 2.00 m/s

**Table 3.**
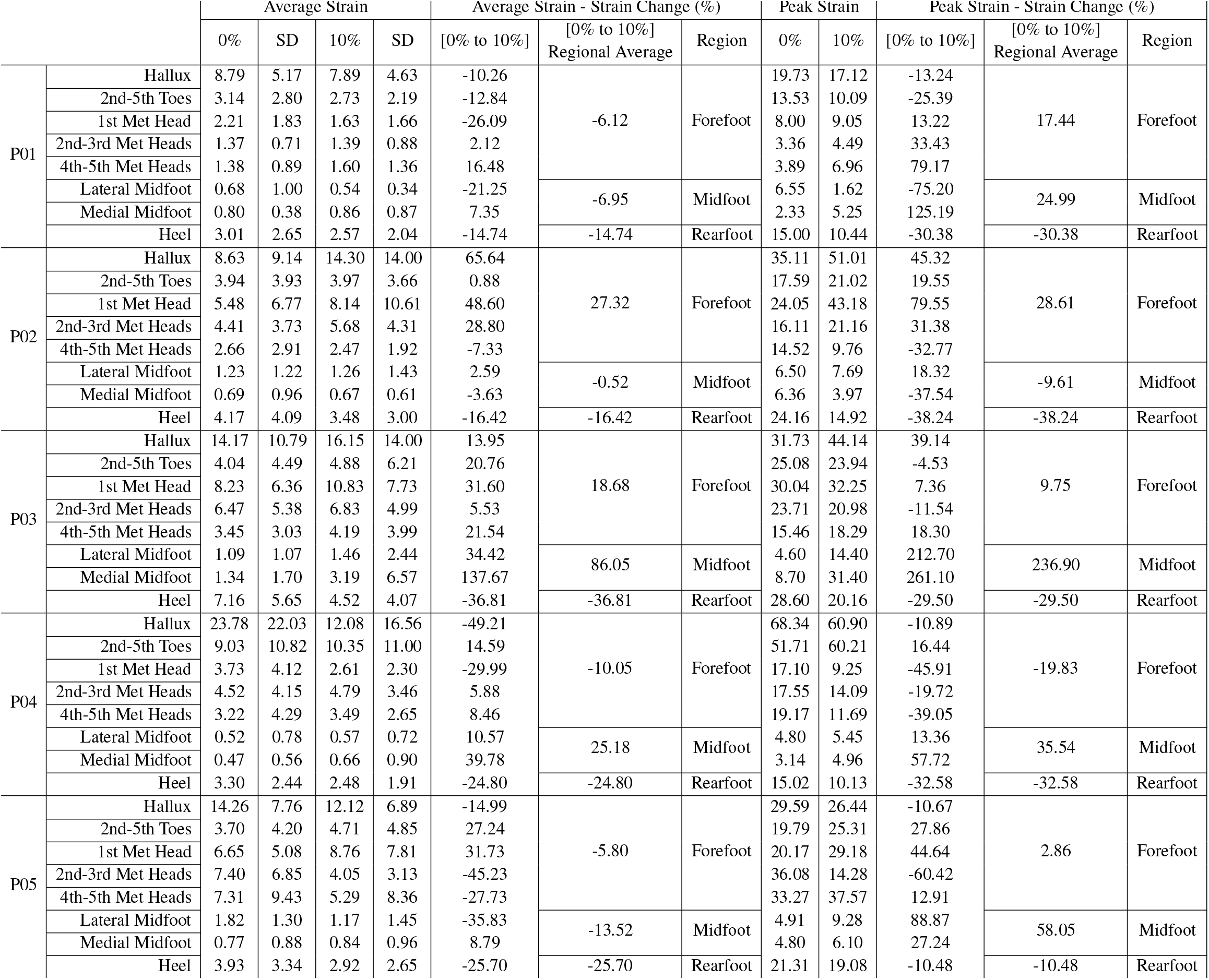
Anatomical regional average and peak strains to 2 d.p. and standard deviations (SD) averaged across all three repeat trials, with comparative percentage strain changes for all participants (P01-P05) at 1.25 m/s speed at 0 % and 10% inclination.

Average stance time, across all ten stance phases and over three repeated trials per loading regime (Fig4). decreases with increasing speed for all participants. A marginal decrease in average stance time is seen for all participants comparative between 1.25 m/s 0% to 1.25 m/s 10% inclination.

**Figure 4.**
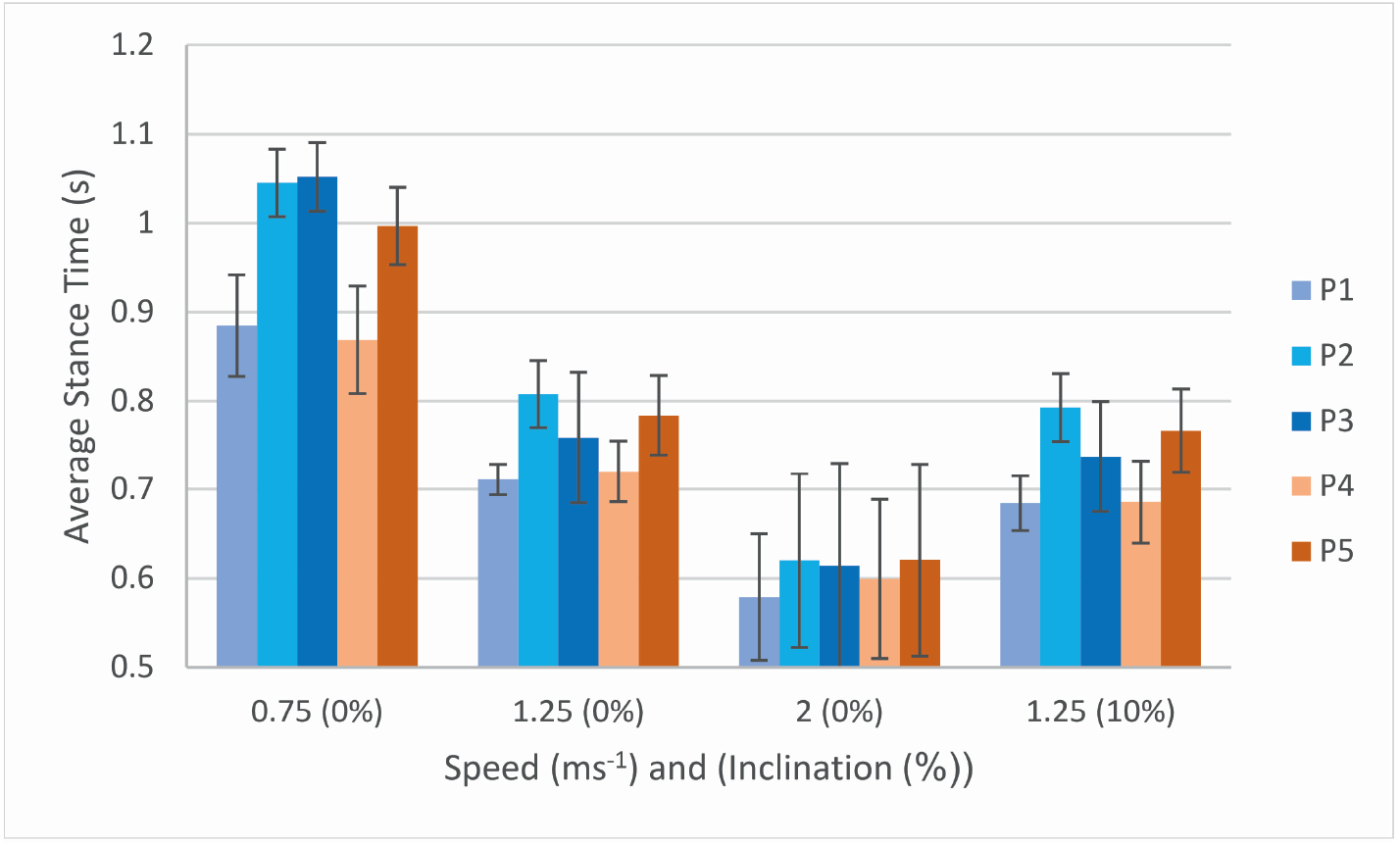
Average stance time per participant across all plantar loading regimes with associated standard deviations.

## 4 DISCUSSION

The aim of the study was to utilise the STAMPS novel measurement insole to evaluate strain characteristic changes (Crossland et al., 2023), including stance time, strain location and strain change, instigated through changes in walking speed and inclination. The strain responses captured using the STAMPS insole were compared to the capabilities of current pressure measurement systems used in DFU assessment, namely the pedar® [Novel GmbH, Munchen Germany] pressure capture insole. Studies by Segal et al. (2004) and Ho et al. (2010) using the pedar® insole showed increased pressure with increasing speed. Strain captured by the STAMPS insole is related to plantar loading comprised of pressure and shear strain contributions, it is therefore expected that with an increasing speed and associated pressure, an increase in strain would be observed.

Strain outputs for all participants, Table 2, confirm this expectation by showing increased strain consistently across all trials with increasing speed (Segal et al., 2004). Average stance time, Fig. 4, reduces with increasing speed which is concurrent with expectations for normal gait (Kirtley et al., 1985; Roth et al., 1997; Demur and Demura, 2010; Olney et al., 1994). Inclination strain change reductions across participants for both average and peak strains, Table 3, align with the reduction in rearfoot strain active locations seen, example shown in Fig. 3. Though there is variation in participant strain changes to the mid and forefoot in both average and peak strain, the peak strain changes in these regions tend to show a general increase for participants at 10% gradient. Ho et al. (2010) reported the effect of inclination on pressure showed reduction in peak pressures at the rearfoot at inclinations of 5%, 10% and 15% gradient, with changes to the location of strain actives regions present from the 0% gradient. This is congruent with the strain changes reported from this study. The gradient chosen for this study does not necessarily reflect the daily gait activities of a person at-risk of diabetic foot complications. A 10% gradient can be considered a moderate-large incline, and with high-risk diabetics skewing towards and older and less mobile population, their daily activity locations may be centred around lower gradient terrains such as the home. Further studies at a reduced gradient and during other activities such as stair ascending and descending would be beneficial to understand strain response variations.

The regional strain percentage decreases reported for some participants across the trialled conditions for speed and gradient do not necessarily reflect the strain changes across the foot as a whole. While a tendency for the whole foot to show increasing strain with increasing speed is seen, the decreasing regional values may reflect the loading changes required of the foot to offset gait deviations undertaken in response to increasing speed as seen in Kernozek et al. (1996) shod pressure study. Likewise the same can be seen in the response of the foot with inclination reported by Ho et al. (2010).

Peak plantar pressure has been considered in DFU analysis alongside pressure time integral as metric used to determine DFU risk (Bus and Waaijman, 2013; Waaijman and Bus, 2012; Keijsers et al., 2010). Whilst the STAMPS insole does not allow for the recording of strain changes during stance phase for direct calculation of strain time integral, and instead provides a reflection of cumulative strain, analysis of regional average strain in relation to longevity of stance can be considered in lieu of this metric. The small cohort in this study does not allow for statistical analysis and reporting of significance, but can be considered a benchmark study to assess this metric in a larger healthy cohort.

A limitation of this study is the use of a treadmill to standardise the walking speeds achieved. Whilst it achieves that aim, it can result in altered gait biomechanics compared to non-treadmill walking to compensate for controlled speed and belt movement (Lee and Hidler, 2008). Therefore the strain profiles may be altered in comparison to strain results recorded due to natural speed changes. A study analysing the strain response of self-selected slow and faster walking speeds should be run to address this.

The acceleration profile of the belt was dependent on the target outcome speed, with lower speeds having a lower initial rate of acceleration comparative to the higher target speed. The acceleration profiles were also non-liner in presentation. Speeds in all three targeted trials were reached prior to half of the steps being completed in all cases, with the plastically deformable STAMPS insole recording peak strains occurring at the target speed. The acceleration profile of the treadmill, rather than immediate target speed reached, does enable the full ten steps undertaken in the trial to be conducted at the target speed. The increased time it takes to reach higher speeds, due to the same initial starting speed, means that a differing number of steps are completed at the target velocity in relation to which speed the trial was conducted at. The result this has on the strain outcomes should be minimal due the plastic deformation of the insole, however this cannot be measured in the remit of this study. Acceleration profiles can be present when achieving self-selected walking speeds within brisk activities of daily living. These acceleration profiles are often not seen with slower self-selected walking speeds, which can be achieved instantaneously from initiation of gait. These natural deviations from a single continuous walking speed, whilst not directly represented in the study due to the controlled acceleration profile, should be considered when assessing the feasibility of strain data capture methods to respond to change in speed during gait events.

The participant study was conducted with a small cohort of healthy participants to assess both if there was a difference seen in strain response and stance time, and if this was measurable using the STAMPS insole technique. Due to this, no statistical significance can be attributed to the strain differences reported, a larger cohort study is required with appropriate power to further this work. To work towards translation of the data to reflect a range of activities of daily living in clinical decision making process, a range of inclinations should be trialled over a larger population.

A singular inclination value is studied at a relatively steep incline of 10%, to report how inclination affects strain response. Beyond this the cohort demographic only covers a young adult, non-diabetic population, meaning that it is not generalizable to other cohorts. However the opportunity to measure strain and potentially reduce the incidence of DFU requires further studies in this population. Development of 3D

DIC image capture is also required to enhance the analysis of the insole deformation profiles, ensuring the recorded strains reflect a true representation of regionalised strain response. This is particularly important in relation to potential future clinical translation to allow for strain data to support DFU risk assessment and treatment pathways.

Prior use of the insole has been limited to normal self-selected walking speeds to reflect the average patients gait speed undergoing activities of daily living, but expanding this to reflect the altered activities of daily living experienced by diabetic cohorts due to foot structure, deformities and gait deviations is important. Assessment of the characteristics of these daily activities has been increasing in recent years and emphasises the importance of characterising gait beyond a research setting (Rozema et al., 1996). Understanding strain response to speed and inclination offers the opportunity to provide informed treatment approaches, such as footwear design, to optimise plantar loading for reduced DFU risk. With the increase in biomechanical assessments of activities of daily living, the clinical translation potential of the STAMPS insole could also be optimised to explore pathology and disease progression through plantar loading.

## Data Availability

All data produced are available online at https://doi.org/10.5518/1309
[DOI due to go live shortly]

## ACKNOWLEDGMENTS

I would like to extend my thanks to Leeds Teaching Hospitals NHS Trust for allowing me to work in their Diabetic Limb Salvage Service on an honorary basis which has influenced my drivers and motivation for this study. Thanks should also be extended to my supervision team for their continual support and guidance and to EPSRC for funding this research through the CDT Centre Grant EP/L01629X/1.

## AUTHOR CONTRIBUTIONS

SRC: Writing – original draft, project administration, methodology, investigation, formal analysis, data curation and conceptualisation. HJS: Writing – review and editing, conceptualisation. CLB: Writing – review and editing, supervision, methodology, conceptualisation PC: Writing – review and editing, supervision, methodology, conceptualisation.

## CONTRIBUTION TO THE FIELD STATEMENT

The research reported in this study adds to a growing body of literature addressing and enhancing the risk assessment methodologies of the vulnerable diabetic foot. The focus of current methods is centred on pressure assessment whilst neglecting the contribution of shear in the formation of diabetic foot ulcers, which are a leading cause of non-traumatic amputations. This research assesses a novel strain measurement insole, detecting the combined contributions of pressure and shear, and looks to assess the strain response changes brought about by changes in speed and inclination. Current risk assessment is often currently conducted within the clinical environment, with an emerging focus on data collection during more representative activities of daily living taking focus. Understanding strain response, as detailed in this study, during these daily activities, such as when changing speed, is important to be able to work towards characterising at-risk plantar sites within the diabetic population.

## ETHICS

Ethics reference: LTMECH-005. University of Leeds Engineering and Physical Sciences joint Faculty Research Ethics Committee provided ethics approval. Prior consent was obtained from all participants within the study.

## SUPPLEMENTAL DATA

Supplementary figures are provided as an additional document showing the remaining participant strain profiles for repeated trialled speeds and inclinations.

## DATA AVAILABILITY STATEMENT

(Currently in process of getting DOI from Leeds Data Repository)

## REFERENCES

Abdul Razak, A. H., Zayegh, A., Begg, R. K., and Wahab, Y. (2012). Foot plantar pressure measurement system: A review. Sensors (Switzerland) 12, 9884–9912. doi:10.3390/s120709884

Armstrong, D. G., Boulton, A. J., and Bus, S. A. (2017). Diabetic Foot Ulcers and Their Recurrence. New England Journal of Medicine 376, 2367–2375. doi:10.1056/NEJMra1615439. Publisher: Massachusetts Medical Society

Bus, S. A., Lavery, L. A., Monteiro-Soares, M., Rasmussen, A., Raspovic, A., Sacco, I. C. C., et al. (2020). Guidelines on the prevention of foot ulcers in persons with diabetes (IWGDF 2019 update). Diabetes/Metabolism Research and Reviews 36. Publisher: John Wiley and Sons Ltd

Bus, S. A. and Waaijman, R. (2013). The value of reporting pressure–time integral data in addition to peak pressure data in studies on the diabetic foot: A systematic review. Clinical Biomechanics 28, 117–121. doi:10.1016/J.CLINBIOMECH.2012.12.002. Publisher: Elsevier

Chatwin, K. E., Abbott, C. A., Rajbhandari, S. M., Reddy, P. N., Bowling, F. L., Boulton, A. J. M., et al. (2021). An intelligent insole system with personalised digital feedback reduces foot pressures during daily life: An 18-month randomised controlled trial. Diabetes Research and Clinical Practice 181, 109091. doi:10.1016/j.diabres.2021.109091

Chijiiwa, K., Hatamura, Y., and Hasegawa, N. (1981). Characteristics of Plasticine Used in the Simulation of Slab in Rolling and Continuous Casting. Transactions of the Iron and Steel Institute of Japan 21, 178–186. doi:10.2355/isijinternational1966.21.178

[Dataset] Crossland, S. R., Jones, A. R., Nixon, J. E., Siddle, H. J., Russell, D. A., and Culmer, P. R. (2023). enSTrain Analysis and Mapping of the Plantar Surface (STAMPS) – A novel technique of plantar load analysis during gait. doi:10.1101/2023.03.10.23287086. Pages: 2023.03.10.23287086

Demur, T. and Demura, S.-i. (2010). Relationship among Gait Parameters while Walking with Varying Loads. Journal of PHYSIOLOGICAL ANTHROPOLOGY 29, 29–34. doi:10.2114/jpa2.29.29

Heuch, L. and Streak Gomersall, J. (2016). Effectiveness of offloading methods in preventing primary diabetic foot ulcers in adults with diabetes: a systematic review. JBI Evidence Synthesis 14, 236. doi:10.11124/JBISRIR-2016-003013

Ho, I.-J., Hou, Y.-Y., Yang, C.-H., Wu, W.-L., Chen, S.-K., and Guo, L.-Y. (2010). Comparison of Plantar Pressure Distribution between Different Speed and Incline During Treadmill Jogging. Journal of Sports Science & Medicine 9, 154–160

International Diabetes Federation (2019). IDF Diabetes Atlas 9th Edition. Tech. rep., International Diabetes Federation, International

Jones, A. D., De Siqueira, J., Nixon, J. E., Siddle, H. J., Culmer, P. R., and Russell, D. A. (2022). engPlantar shear stress in the diabetic foot: A systematic review and meta-analysis. Diabetic Medicine: A Journal of the British Diabetic Association 39, e14661. doi:10.1111/dme.14661

Keijsers, N. L. W., Stolwijk, N. M., and Pataky, T. C. (2010). Linear dependence of peak, mean, and pressure–time integral values in plantar pressure images. Gait & Posture 31, 140–142. doi:10.1016/j.gaitpost.2009.08.248

Kernozek, T., LaMott, E., and Dancisak, M. (1996). Reliability of an In-Shoe Pressure Measurement System During Treadmill Walking. Foot & ankle international / American Orthopaedic Foot and Ankle Society [and] Swiss Foot and Ankle Society 17, 204–9. doi:10.1177/107110079601700404

Kerr, M., Barron, E., Chadwick, P., Evans, T., Kong, W. M., Rayman, G., et al. (2019). The cost of diabetic foot ulcers and amputations to the National Health Service in England. Diabetic Medicine 36, 995–1002. doi:10.1111/dme.13973. Publisher: Blackwell Publishing Ltd

Kirtley, C., Whittle, M. W., and Jefferson, R. J. (1985). Influence of walking speed on gait parameters. Journal of Biomedical Engineering 7, 282–288. doi:10.1016/0141-5425(85)90055-X

Lavery, L. A., Armstrong, D. G., Wunderlich, R. P., Tredwell, J., and Boulton, A. J. (2003). Predictive value of foot pressure assessment as part of a population-based diabetes disease management program. Diabetes care 26, 1069–1073. doi:10.2337/DIACARE.26.4.1069. Publisher: Diabetes Care

Lee, S. J. and Hidler, J. (2008). Biomechanics of overground vs. treadmill walking in healthy individuals. Journal of Applied Physiology 104, 747–755. doi:10.1152/japplphysiol.01380.2006. Publisher: American Physiological Society

Lord, M. and Hosein, R. (2000). A study of in-shoe plantar shear in patients with diabetic neuropathy. Clinical Biomechanics 15, 278–283. ISBN: 02680033/00

Michael A., M. A., Orteu, J.-J., and Schreier, H. W. (2009). Digital Image Correlation (DIC). In Image Correlation for Shape, Motion and Deformation Measurements (Boston, MA: Springer US). 1–37. doi:10.1007/978-0-387-78747-35

Olney, S. J., Griffin, M. P., and McBride, I. D. (1994). Temporal, Kinematic, and Kinetic Variables Related to Gait Speed in Subjects With Hemiplegia: A Regression Approach. Physical Therapy 74, 872–885. doi:10.1093/ptj/74.9.872

Padulo, J., Chamari, K., and Ardigò, L. P. (2014). Walking and running on treadmill: the standard criteria for kinematics studies. Muscles, Ligaments and Tendons Journal 4, 159–162

Pit’hová, P., Pátková, H., Galandáková, I., Dolezalová, L., and Kvapil, M. (2007). Differences in ulcer location in diabetic foot syndrome. Vnitrni Lekarstvi 53, 1278–1285

Rajala, S. and Lekkala, J. (2014). Plantar shear stress measurements - A review. Clinical Biomechanics 29, 475–483. doi:10.1016/j.clinbiomech.2014.04.009. Publisher: Elsevier Ltd

Roth, E. J., Merbitz, C., Mroczek, K., Dugan, S. A., and Suh, W. W. (1997). Hemiplegic gait. Relationships between walking speed and other temporal parameters. American Journal of Physical Medicine & Rehabilitation 76, 128–133. doi:10.1097/00002060-199703000-00008

Rozema, A., Ulbrecht, J. S., Pammer, S. E., and Cavanagh, P. R. (1996). In-Shoe Plantar Pressures During Activities of Daily Living: Implications for Therapeutic Footwear Design. Foot & Ankle International 17, 352–359. doi:10.1177/107110079601700611. Publisher: SAGE Publications Inc

Segal, A., Rohr, E., Orendurff, M., Shofer, J., O’Brien, M., and Sangeorzan, B. (2004). The Effect of Walking Speed on Peak Plantar Pressure. Foot & Ankle International 25, 926–933. doi:10.1177/107110070402501215. Publisher: SAGE Publications Inc

van Netten, J. J., van Baal, J. G., Bril, A., Wissink, M., and Bus, S. A. (2018). engAn exploratory study on differences in cumulative plantar tissue stress between healing and non-healing plantar neuropathic diabetic foot ulcers. Clinical Biomechanics (Bristol, Avon) 53, 86–92. doi:10.1016/j.clinbiomech.2018.02.012

Waaijman, R. and Bus, S. A. (2012). The interdependency of peak pressure and pressure–time integral in pressure studies on diabetic footwear: No need to report both parameters. Gait & Posture 35, 1–5. doi:10.1016/j.gaitpost.2011.07.006

Warren, G. L., Maher, R. M., and Higbie, E. J. (2004). Temporal patterns of plantar pressures and lower-leg muscle activity during walking: effect of speed. Gait & Posture 19, 91–100. doi:10.1016/S0966-6362(03)00031-6

Yavuz, M., Botek, G., and Davis, B. L. (2007). Plantar shear stress distributions: Comparing actual and predicted frictional forces at the foot–ground interface. Journal of Biomechanics 40, 3045–3049. doi:10.1016/j.jbiomech.2007.02.006

Yavuz, M., Tajaddini, A., Botek, G., and Davis, B. L. (2008). Temporal characteristics of plantar shear distribution: relevance to diabetic patients. Journal of Biomechanics 41, 556–559. doi:10.1016/j.jbiomech.2007.10.008

