## Supplementary Figures for "Evaluating the use of a novel low-cost measurement insole to characterise plantar foot strain during gait loading regimes"

### Supplementary Material

#### 1 SUPPLEMENTARY FIGURES

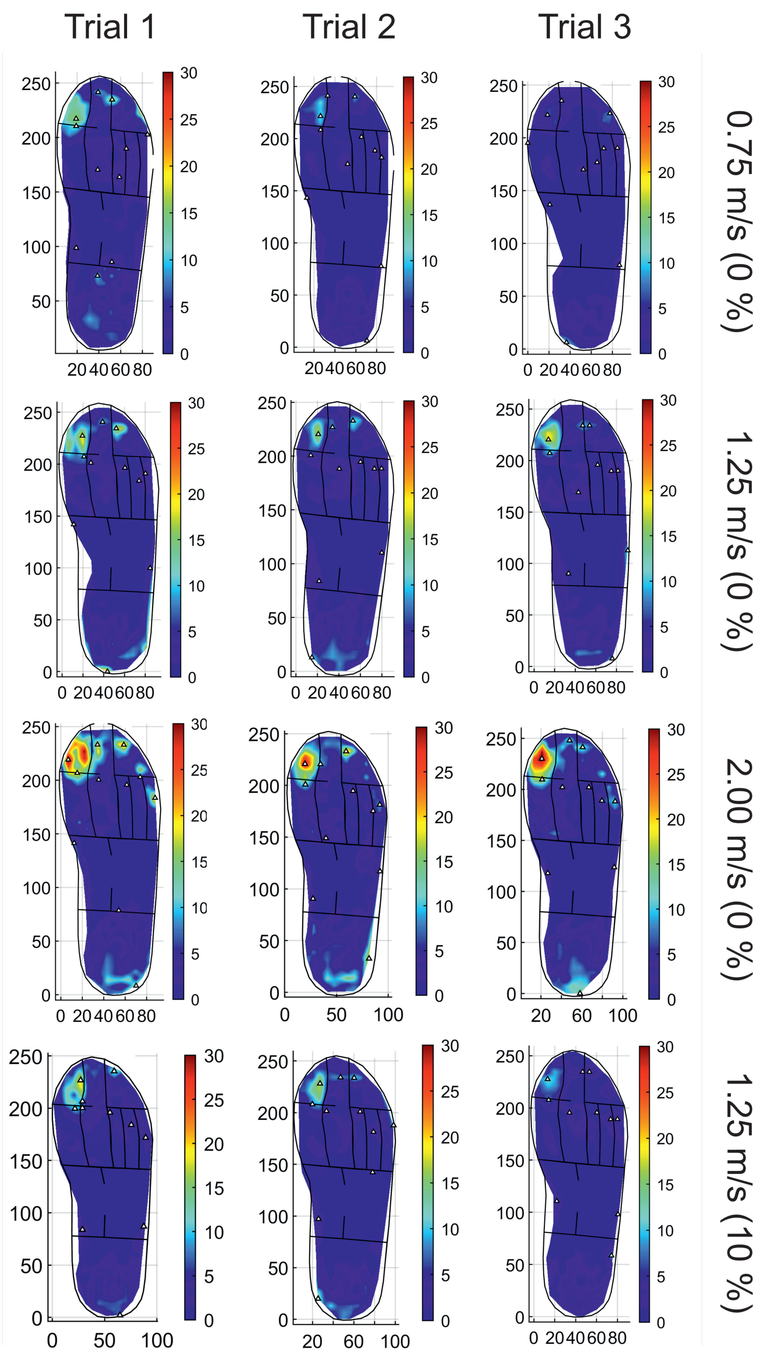


**Figure S1.** Strain profiles for repeated trialled speeds and inclinations for P01.

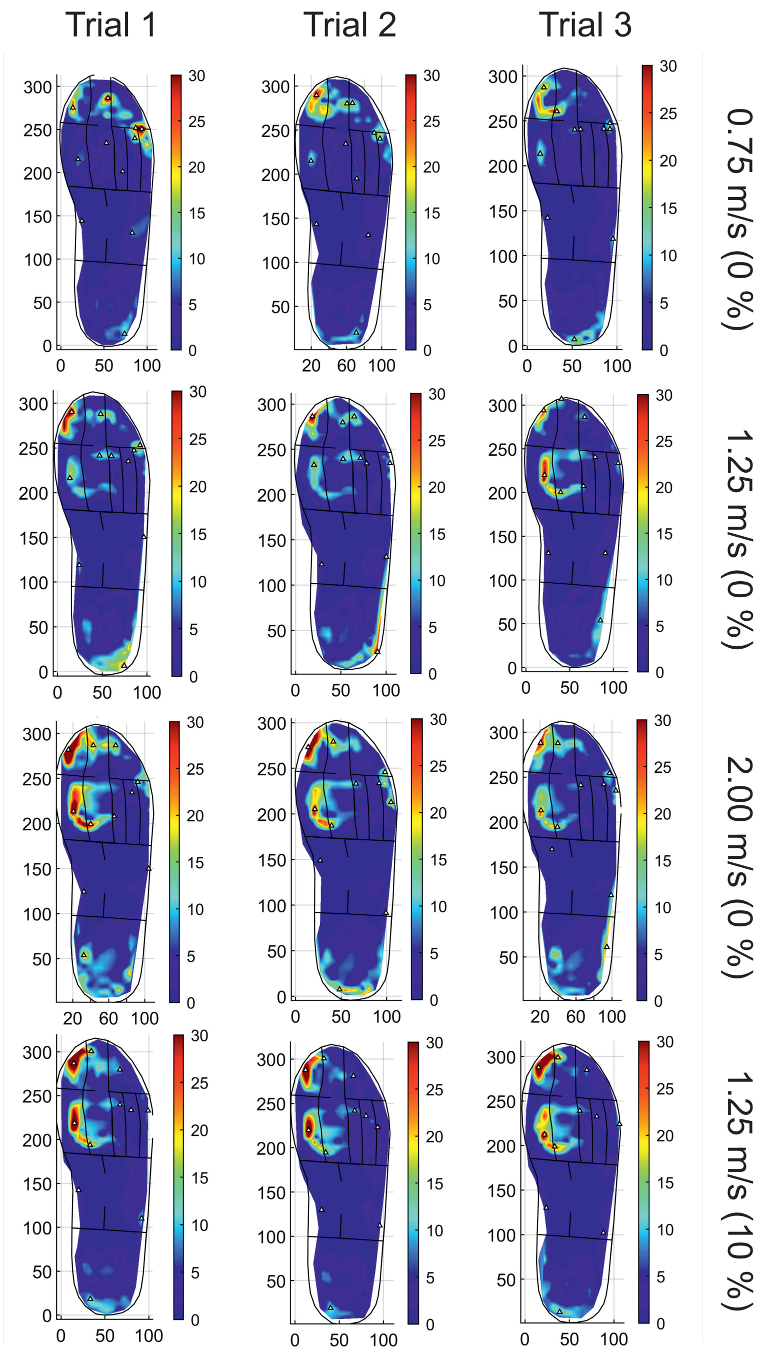


**Figure S2.** Strain profiles for repeated trialled speeds and inclinations for P02.

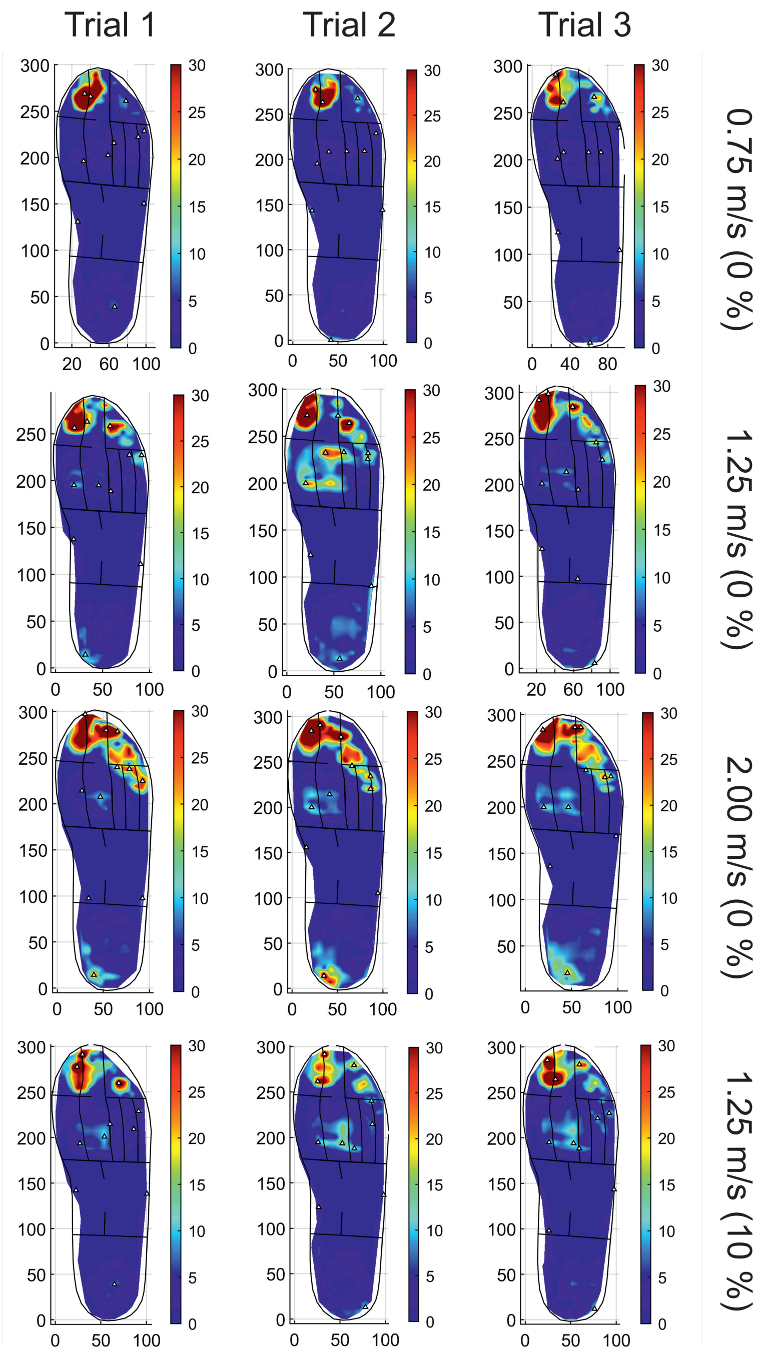


**Figure S3.** Strain profiles for repeated trialled speeds and inclinations for P04.

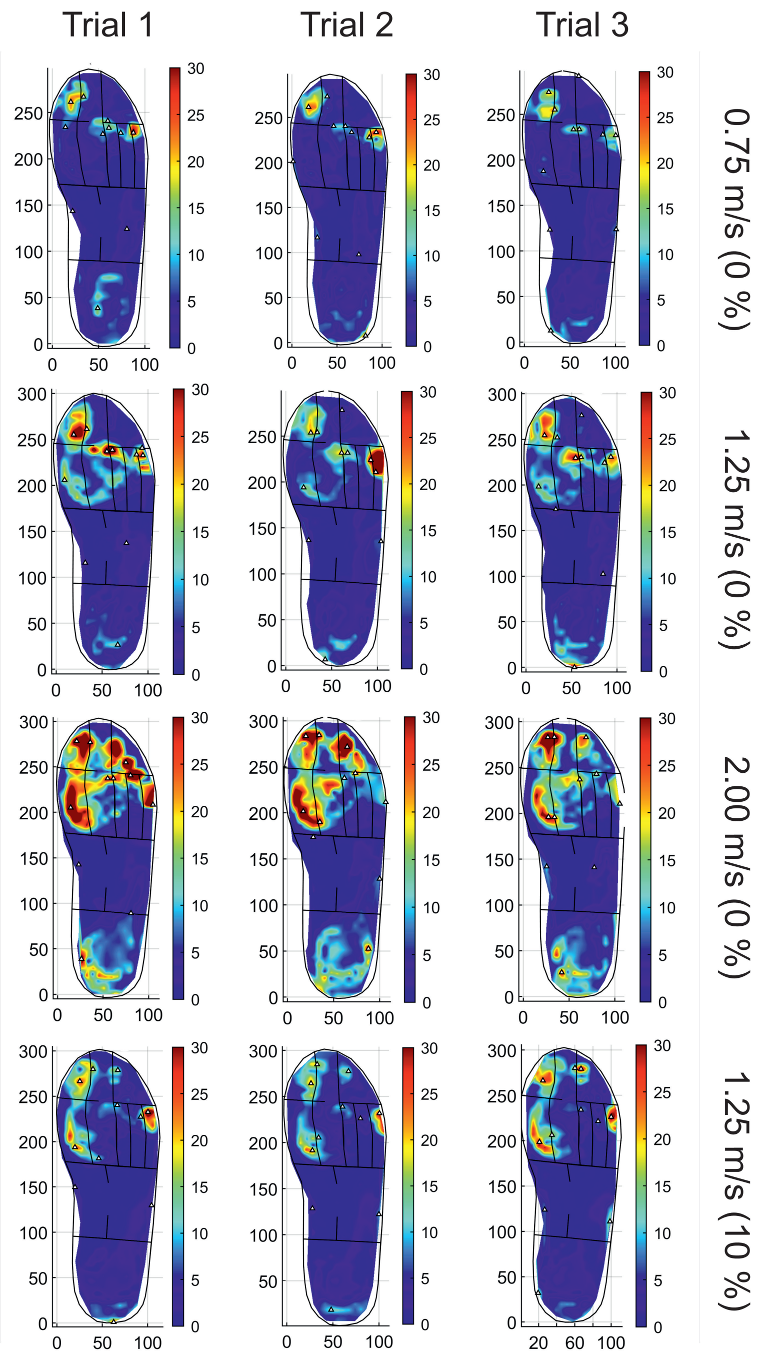


**Figure S4.** Strain profiles for repeated trialled speeds and inclinations for P05.
